# Optimal deployment of limited vaccine supplies to combat mpox

**DOI:** 10.1101/2024.11.03.24316551

**Authors:** Matthew T Berry, C Raina MacIntyre, Deborah Cromer, Adam Hacker, Miles P Davenport, David S Khoury

## Abstract

Mpox outbreaks in Central Africa have been declared a public health emergency of international concern by the World Health Organization. Fortunately, real-world effectiveness studies of the MVA-BN vaccine indicate that it has an effectiveness of 74% after one dose, and 82% after two doses against mpox. However, given the very limited supply of vaccines in Central Africa, there remain questions around the optimal deployment of limited MVA-BN doses. In this study, we consider whether more mpox cases might be averted by following the traditional two-dose vaccine regimen (4 week dosing interval), or by giving a single dose of MVA-BN to as many individuals as possible. We find that the optimal strategy depends on both, (i) the degree to which a subpopulation might be at higher risk of mpox, or severe mpox, infections, and (ii) how long ago the first dose was administered to the most at-risk subpopulation.

## Introduction

There are concurrent outbreaks of mpox clades I and II globally^1^. Of great concern is the current clade I outbreak in central Africa with high case rates, increasing transmission and case-fatality ratio reported at approximately 2% (African continent average), and higher in children^2,3^. Fortunately, there is considerable evidence that vaccinia-based vaccines are effective at preventing both mpox infection and at reducing the severity of illness^4–6^. The evidence of vaccine effectiveness (VE) is derived from real-world effectiveness studies of the third generation MVA-BN vaccine in western populations during the clade IIb pandemic of 2022-2023^7–10^. A number of older studies in the Democratic Republic of Congo (DRC, formally Zaire) have also shown that vaccination with first generation vaccinia-based vaccines is protective against mpox^11–13^. However, third generation vaccines are preferable to early vaccinia-vaccines because of the superior safety and utility in immunosuppressed individuals^14^. Despite the urgent need to vaccinate at-risk populations, particularly in the DRC and neighbouring countries, vaccine availability is limited^15^. Africa CDC estimate 10 million doses are required to control the epidemic^16^. Japan has committed 3 million doses of LC16, although to date no supply has reached DRC or neighbouring countries.

Approximately 200,000 doses of MVA-BN (Jynneos) arrived in DRC in September 2024. LC16 is a single dose regimen, while MVA-BN is currently recommended as a two-dose regimen (given 4 weeks apart)^17^. The DRC has a population of more than 110 million.

Therefore, how to optimally deploy a limited stock of vaccines to avert the greatest number of clinical cases, particularly severe cases, is a critical question at present.

Recently, multiple meta-analyses of real-world effectiveness studies of MVA-BN have analysed the effectiveness of one-dose and two-dose MVA-BN against clade IIb mpox^4–6^. These have shown a relatively incremental improvement in vaccine effectiveness after two doses compared with a single dose. For example, the World Health Organization (WHO) states that one dose and two doses of MVA-BN provide 76% and 82% effectiveness against mpox, respectively^18^. In our own meta-analysis we estimated very similar vaccine effectiveness of 74% after one dose and 82% after two doses. Based on this comparison, we commented that in the case of limited vaccine supply, it would be optimal to deploy a single dose of vaccine to as many individuals as possible rather than administer two doses to half as many individuals^4^. However, this analysis did not include modelling of the waning of vaccine effectiveness with time after one dose vaccination (i.e. the comparison was based on reported vaccine effectiveness from clinical studies, and did not model the predicted decay of vaccine effectiveness over time). Further, the analysis only considered the situation of a population at homogeneous risk of infection, but did not consider the implications of high-risk groups for both infection and for severe outcomes, as is currently observed across age groups in the clade I outbreak in the DRC^2,19^. Here we consider the question of the optimal deployment of a limited number of MVA-BN doses given the likely waning of vaccine effectiveness, and in a population where there are subgroups at different risk of infection and severe outcomes. In particular, we are interested in the decision, at a particular time point, of whether to use a limited supply of MVA-BN vaccines to deliver a second dose to those who have already received a first dose of MVA-BN or to deploy a first dose of the limited vaccine stock to naïve individuals. This question does not consider the reason individuals may have received a first dose in the past (e.g. either as post-exposure prophylaxis or as primary prevention), or the protection the first vaccine doses have provided up to the time our decision must be made. But rather, we only ask, from the time of our current decision on deployment, what is the relative advantage of deploying a limited stock of these vaccines as second doses to those already partially vaccinated or as first doses to those who have not yet received any vaccine doses?

It is important to note that, in this study, we rely on data and modelling of MVA-BN effectiveness derived from the clade IIb global outbreaks, but throughout our analysis we discuss these results assuming they are translatable to the contemporaneous clade I outbreaks in Africa. Although it is generally thought that these vaccines will also be effective against clade I mpox (since immunity to orthopoxviruses is often cross-protective^14^), confirmatory studies showing that these vaccines are effective against clade I mpox are currently lacking, and are urgently needed. Further, in this analysis we do not explicitly model mpox transmission or consider the impact of different vaccination strategies on transmission or the epidemic trajectory, since there is limited data to inform these outcomes. Instead, we consider the distribution of a limited supply of vaccines where the primary goal is to reduce risk of infection and severe outcomes in particular cohorts.

## Results

### Comparing one-dose versus two-dose MVA-BN vaccination strategies over two years

In a previous study we estimated that distributing limited doses of MVA-BN as a single dose regimen would avert around 1.8-fold more cases of mpox than deploying the same number of doses as a two-dose regimen^4^. This was based only on the comparative reported effectiveness of the one and two dose regimens (fig. 1A). However, vaccinia-binding antibody titres wane over time, and if antibody responses are predictive of protection, vaccine effectiveness against symptomatic infection may wane in parallel^4^ (fig. 1B). Therefore, in this case, it is important to model the possible effect of this decay on the comparative benefits of a one-vs. two-dose regimen. We consider a scenario where one dose of vaccine has been deployed to some groups of individuals either as primary prevention or post-exposure prophylaxis, and at some later timepoint authorities will have a choice to either administer a second dose to those same individuals already primed, or to administer a first dose of vaccine to naïve individuals (fig. 1C). Without modelling the effects of waning immunity, using only the estimates of vaccine effectiveness from our previous meta-regression of real world effectiveness studies^4^, a one-dose vaccination schedule is predicted to avert 1.80-fold (CI: 1.50-1.92), as we reported previously. In order to account for the impact of waning vaccine effectiveness, assuming a fixed force of infection over time, the ratio of the cases averted with each strategy can be calculated as the ratio of the average vaccine effectiveness over a given time interval, beginning when the vaccines are deployed under one or the other strategy (note that we also consider the more general case of a varying force of infection in the supplementary material). Modelling of the decay of vaccinia-binding antibody titres and protection suggests that the benefit of administering a first dose to as many people as possible is relatively constant over the first two years after vaccination (fig. 1D). In the subsequent analysis we focus on a time interval of 2 years after deployment, by which time, if an outbreak continues to intensify, vaccine manufacturing and supply are likely to have increased.

**Figure 1:**
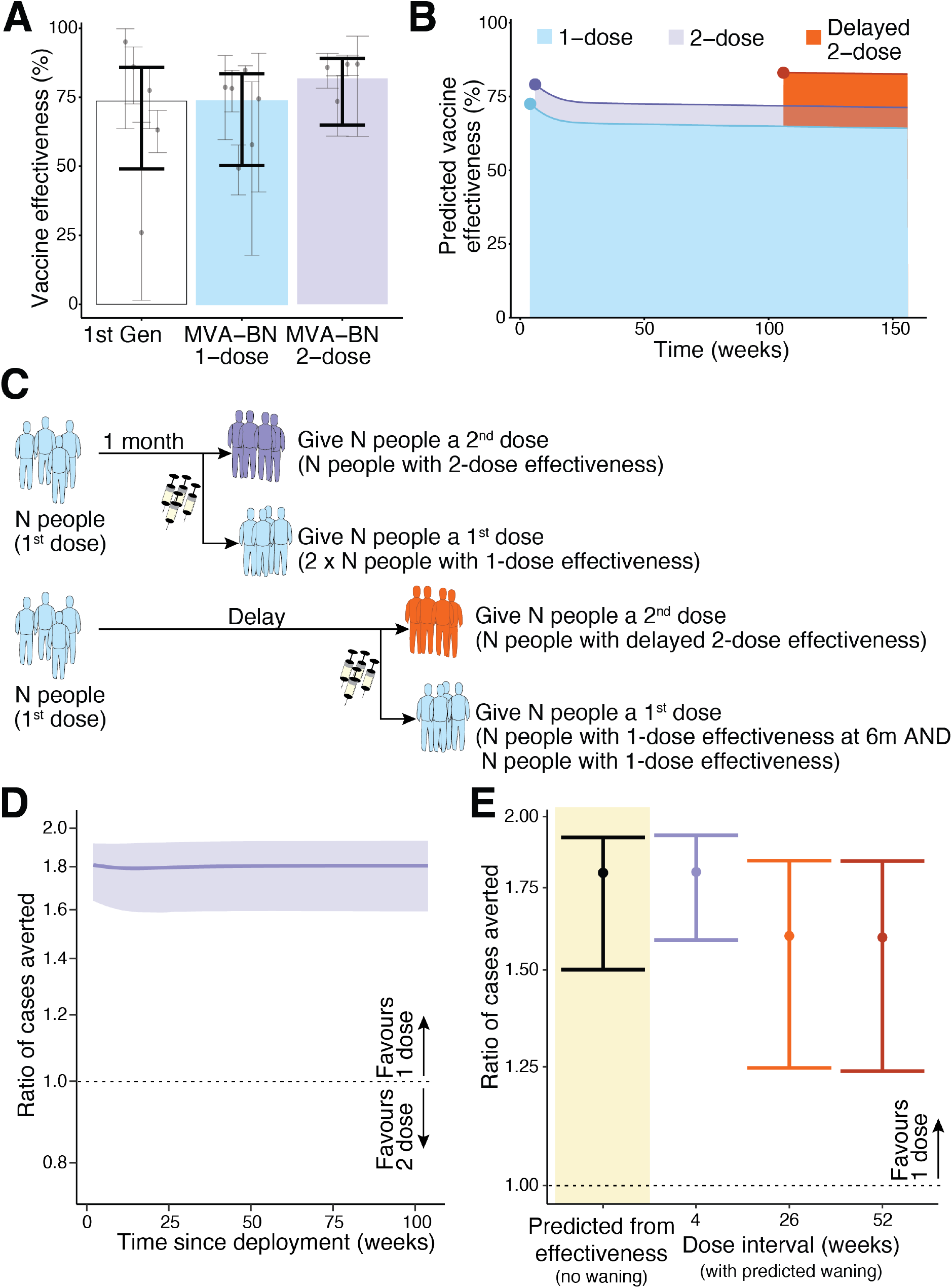
Estimating the protection from using MVA-BN in a one versus two-dose schedule. A) The previously reported vaccine effectiveness of first-generation vaccinia immunisation, and one and two dose MVA-BN vaccination (against clade IIb virus) from a systematic review and meta-analysis of the available data^4^. B) The predicted vaccine effectiveness over time for one dose, two doses spaced 4 weeks apart, and delayed two dose (administered at 2 years). These predictions are from reference ^4^, where a model-based meta-analysis was performed that linked reports of vaccinia-binding antibody titres after MVA-BN vaccination and real-world effectiveness studies, and vaccine effectiveness over time was extrapolated by assuming these binding titres predict vaccine effectiveness (which is not yet confirmed). C) Schematic of potential vaccine allocation options assuming additional vaccine is available 4 weeks after primary vaccination (top) or at some later timepoint (bottom). In each case we compare the outcome of either giving a second dose to those already vaccinated or allocating the additional vaccine as first doses to a naïve population. D) The ratio of cases averted by administering a first dose to naïve individuals compared to a second dose administered at 4 weeks, measured over the first two years after vaccination. The ratio does not vary much over the time interval being considered. E) The ratio of cases averted by giving a one dose regimen rather than a two-dose regimen, estimated using the vaccine effectiveness observed in a meta-analysis of real world effectiveness studies^4^ (left, black) or predicted from a model-based meta-analysis using predictions of waning and different dose spacing^4^. Note that in the case of giving a second dose at 6 and 12 months we make the assumption that peak antibody binding titres and decay in titres will resemble that seen after giving a second dose at 2 years (the only delayed interval for which immunogenicity data was available^4^). This assumption inflates the predicted benefit of the two-dose regimen.

The analysis above considers the scenario where sufficient vaccine is available to deliver a second dose 4 weeks after the first. However, if vaccine becomes available later after the first dose, does this change the relative benefit of a one-dose vs. a two-dose strategy? Studies have shown that delaying the second dose until 2 years after the first dose leads to a higher peak titre and slower decay of antibody titres post-vaccination^20^. Thus, in addition to declining protection from the initial one-dose vaccination over time, the relative benefit of administering a delayed second dose is expected to increase over time. To model this scenario, we take the same function of waning one-dose immunity as above (fig. 1B). Of note, we only have data on the impact of increasing the spacing of two doses from 1 month to 2 years (fig. 1B), and do not have detailed data on the effects of different dose-spacing intervals on antibody titres. However, we make the assumption that delaying the second dose to six months or more induces a similar peak immune response to that achieved with a second dose at 2 years (fig. 1B), which is an assumption that is expected to overestimate the protection from the two-dose regimen when the second dose is administered at 6 or 12 months. After accounting for waning immunity and higher predicted protection with a delayed second dose, administering a first dose to as many individuals as possible is still predicted to maximise the cases averted from mpox compared to a two-dose strategy (fig.1E). This is observed whether additional vaccine becomes available, 4 weeks, 6 months or 12 months after priming and despite our use of a conservative approach to estimating the effect of a delayed second dose (that will favour a two-dose regimen). We estimate there is still a 1.59-fold (CI: 1.24-1.84) advantage of administering first doses to naïve individuals compared to a strategy of administering a second dose to those already vaccinated individuals at 12 months.

### Considering heterogenous risk groups

In the analysis above we see that in the case of a population at-risk and limited vaccine doses, giving as many people as possible a first dose of MVA-BN would avert more cases than giving half as many people two doses of MVA-BN. However, this only considers a population with homogenous risk of infection and does not consider the possibility of some subgroups being at greater risk of infection and / or of severe outcomes than others. The latest data from the DRC highlights that risk of both infection and the case-fatality ratios of clade I are much higher in young children than older children and adults^2^. Therefore, it is important to consider when it might be optimal to deploy a second dose of MVA-BN to those most at risk, rather than to give a first dose to people at a lower risk for primary prevention of mpox. We consider a scenario of two identifiable risk groups, one with high risk, and one with lower risk of mpox infection. Initially, it is favourable to target the high-risk individuals with a first dose. However, when the majority of the high-risk individuals have already received a first dose of MVA-BN, we ask whether it is optimal to administer a second dose to the high-risk group or deliver a first dose to those in the lower risk group (fig. 2A). Specifically, we consider how many times higher the risk would need to be in a population before it becomes optimal to target high-risk individuals with a second dose instead of targeting lower risk individuals with a first dose (which we refer to as a ‘risk-threshold’ above which the two-dose regimen is preferred).

**Figure 2:**
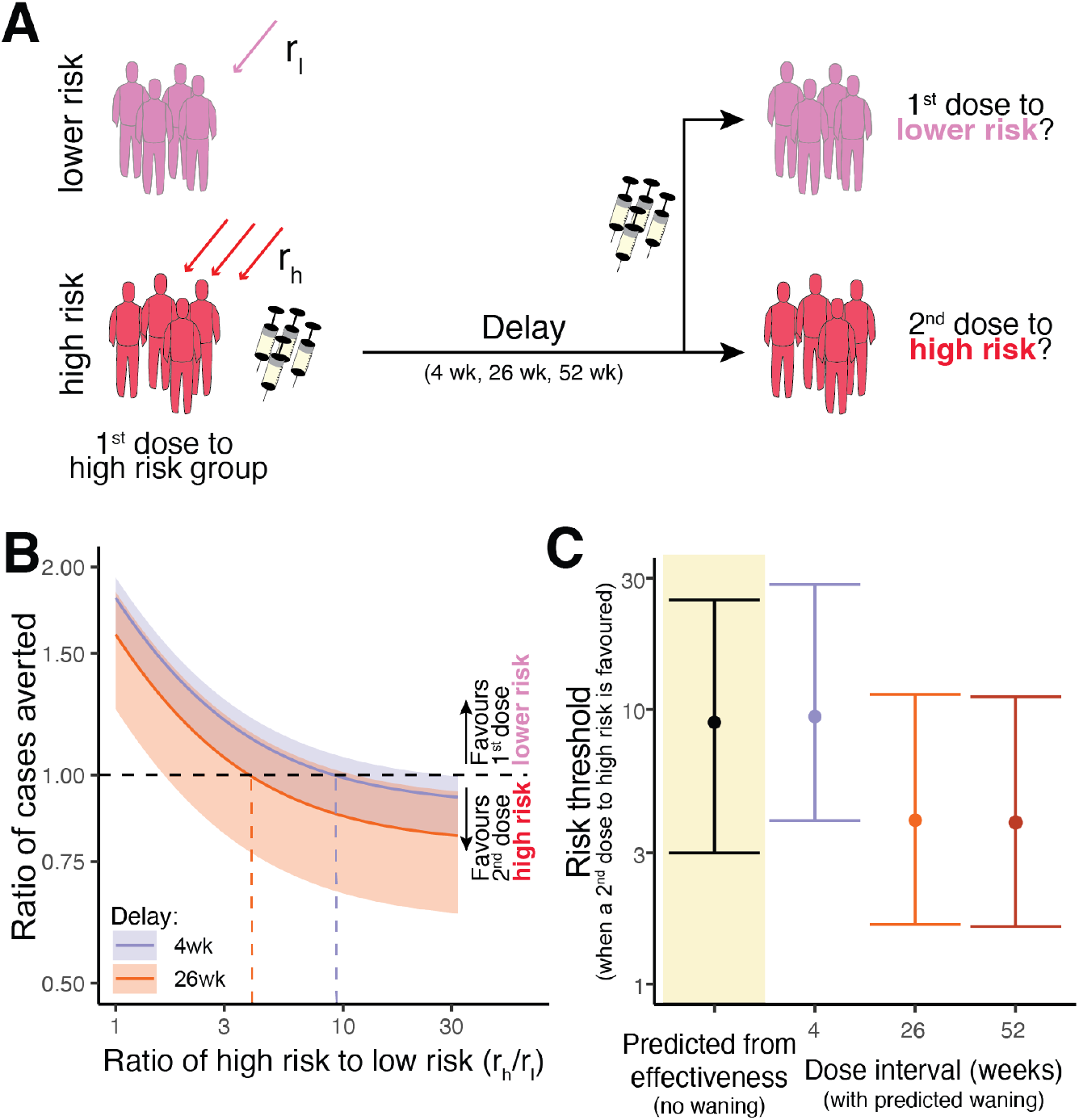
Predicting the effects of distributing vaccine doses between risk groups. A) Schematic of allocation scenario. We assume that a first dose of vaccine has been administered to a high-risk group, and at some later time more vaccine becomes available. These vaccines could either be allocated as second doses to the high-risk group, or administered as a first dose to a lower-risk group. (B) Depending on the ratio of risk in the high-risk and lower-risk groups (x-axis), then the predicted ratio of cases averted by the one-dose compared to the two-dose strategy (y-axis) will vary. This will also vary with the spacing between doses (colours), with longer delays since first dose in the high-risk group favouring the strategy of a second dose to the high-risk group. The horizontal dashed line shows a ratio of one (i.e. where we flip from favouring a one-dose approach to favouring a two-dose approach). Results are shown for prevention of any clinical mpox infection. (C) The risk threshold (y-axis) is the risk ratio at which the optimal strategy changes from favouring a one-dose to favouring a two-dose regimen. The risk threshold predicted from the vaccine effectiveness reported in clinical studies is on the left (black), and the threshold estimated from model-based meta-analysis (including vaccine waning for vaccine administered at 4, 26, or 52 weeks) is shown on the right.

Using only the vaccine effectiveness from our meta-analysis of real-world data, we estimate that unless the high-risk group is at least 9.0-fold (95% CI: 3.0-25) more likely to be infected than the lower risk group, it is preferable to administer any additional vaccines as first doses to the lower risk group. If we take into account waning immunity and protection over the first two years after deployment, we see a similar risk-threshold for when we should favour a two-dose regimen (fig. 2B). If additional vaccines only become available at 6 months or 12 months after the first dose, the risk-threshold is reduced to 3.9-fold (CI: 1.6-11) because of waning immunity after the first dose and a higher response after a delayed second dose (fig. 2C) (using the assumption that a second dose at 6 months or 12 months gives the same peak protection as it would at 2 years, which is a conservative assumption, as it tends to decrease the risk-threshold). Together this suggests that given the relative effectiveness of one vs two doses, unless a subgroup is known to have at least a 3.9-fold (CI:1.6-11) higher risk of infection, it is optimal to continue to administer a first dose to lower risk populations before giving second doses to the high-risk population.

### Averting severe outcomes

In the scenario above we only considered the optimal strategy to avoid the most mpox cases of any severity (mild, moderate or severe). However, case-fatality rates in children under 5 years of age have been reported at 3.2-times higher than in adults and children older than 15 years^2^. These observations are based on very limited epidemiological data and may suffer from a range of confounding^21^. However, if the goal of vaccination were to reduce severe mpox (rather than any clinical mpox infection), this raises the question of whether we should favour deploying limited vaccine stocks as a one- or two-dose regimen, and under what conditions we should target high-risk groups for two-dose vaccination?

Unfortunately, there is very limited data available on vaccine effectiveness in preventing severe mpox infection (and none for clade I infection). However, a comprehensive meta-analysis of studies of individuals with breakthrough mpox clade IIb infection suggested a 66.6% (95% CI: 55-78%) vaccine effectiveness against progression from mild infection to hospitalisation^5^, and there appears similar vaccine effectiveness in preventing progression to severe mpox after either one or two doses of MVA-BN. Assuming that one dose MVA-BN has 74% effectiveness at preventing infection and two doses of MVA-BN has 82% effectiveness, and assuming both provide 66% protection against progressing to severe outcomes, we can estimate from this that the vaccine effectiveness at preventing severe infections that lead to hospitalisation is around 91% for a one-dose, and 94% for a two-dose regimen.

Using only these estimates of vaccine effectiveness from the meta-analysis of real-world studies, we can predict that, for a homogenous population, administering additional vaccines as a first dose to naïve individuals will prevent 1.94-fold (CI: 1.86-1.98) more cases of severe mpox (fig. 3A). We observe similar results when accounting for waning of protection (fig. 3A). Thus, as was the case when looking at averting mpox cases of any severity (fig. 1E), the one-dose strategy is predicted to be optimal if the goal of vaccination is to maximise protection from severe mpox infection.

**Figure 3:**
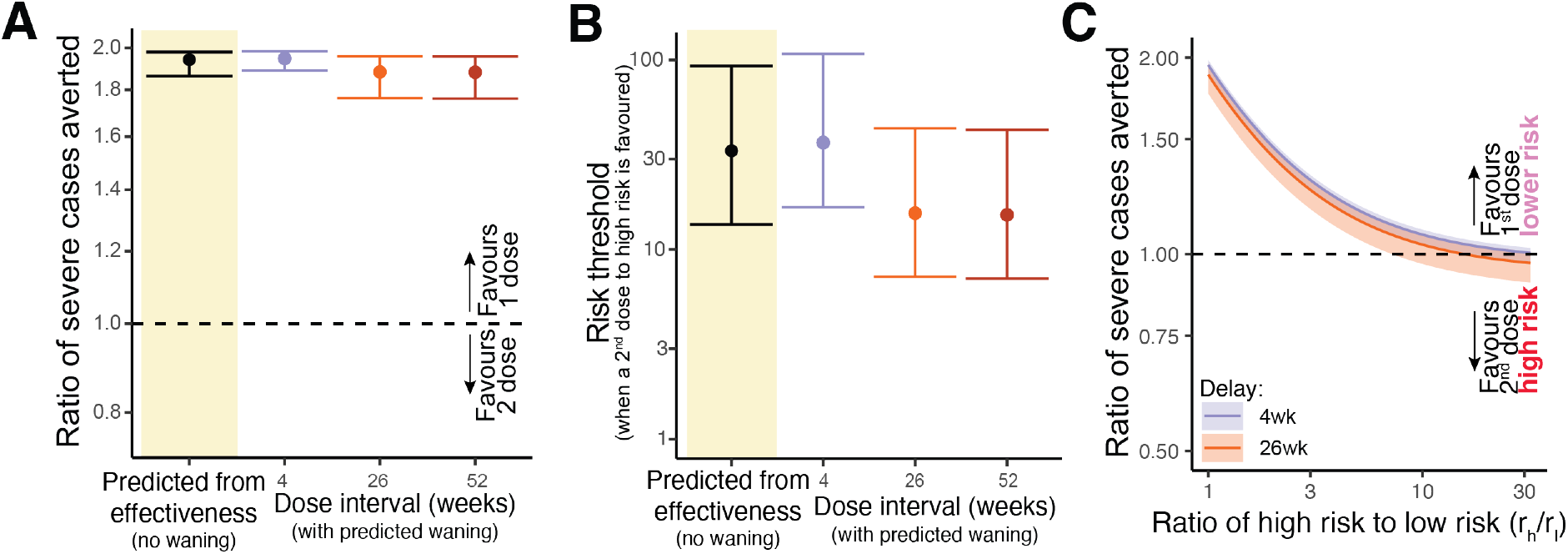
Comparison of vaccine strategies against preventing severe mpox cases. A) The ratio of severe cases averted by giving a one-dose regimen compared to a two-dose regimen - based on the vaccine effectiveness observed in the clinical studies (left, black) or predicted (when the doses are spaced at 4, 26, or 52 weeks) from a model-based meta-analysis (right). B) The risk threshold (the ratio of risk in the high-risk group compared to the low risk) that favours the switch from the one-dose to the two-dose regimen for severe mpox. The risk threshold predicted from the vaccine effectiveness reported in clinical studies (left, black), and the model-based meta-analysis for vaccine administered at 4, 26, or 52 weeks (right). C) The ratio of severe cases averted when comparing vaccination strategies between administering second doses to a high-risk group or first dose to a lower risk group with different dosing intervals (colours). The dashed horizontal line represents the risk threshold where a switch occurs from favouring a one-dose to favouring a two-dose regimen.

As we did above for mpox cases of any severity, we can also analyse the situation where there are identifiable subgroups that are at higher risk of severe infection. Again, here we consider a scenario where the high-risk group has already received one dose of vaccine, and we have the choice of deploying additional vaccine doses as second doses to the high-risk group, or as first doses to a low-risk group. Using only the data from the real-world studies of vaccine effectiveness, we predict that unless the high-risk group were at greater than 33-fold (CI:14-93) higher risk of severe infection than the low-risk group, we should favour administering one dose of the vaccine to as many people as possible rather than giving a second dose to the high-risk population (fig. 3B). The result is similar when accounting for the predicted waning of vaccine effectiveness with time (fig. 3C). That is, applying our predictions of waning vaccine effectiveness^4^, we predict that early after vaccination, giving a first dose to more people would be favoured unless the high-risk group had at least 37-fold (CI:17-107) times higher risk of severe infection compared to the low-risk group (fig. 3B).

Finally, if a second dose is delayed to 6 months or 12 months after the first dose (again using the conservative assumption that a second dose at 6 months or 12 months gives the same peak protection as it would at 2 years), the single dose strategy will remain favoured unless the high-risk group has more than a 15-fold (CI: 7-43) higher risk of severe infection compared to the lower risk group (fig. 3B).

## Discussion

In the context of adequate vaccine availability, delivering the recommended two dose MVA-BN schedule to as many people as possible can achieve rapid population protection. Here we do not advocate for changes to the recommended vaccine schedule generally, or delaying the second dose of MVA-BN as a standard approach. However, in the current mpox public health emergency there are limited vaccine stocks in the countries most in need, which raises a number of questions about the optimal deployment of limited vaccine resources. Here we find that in a population of homogenous risk, deploying limited vaccine stocks as single doses to as many people as possible is always favoured. In the presence of high-risk populations, deploying vaccine as single doses is favoured unless the high-risk population is at greater than 9 times higher risk of clinical mpox infection compared to the low-risk population. For sub-populations at high risk for severe disease, administering one dose to as many people as possible is still favoured unless the high-risk group is at greater than 33-fold higher risk of severe mpox infection (Figure 2E). However, if additional doses of vaccine only become available 6 months to a year after the first dose, the estimated benefit of the one-dose regimen is slightly lower. Waning immunity from the earlier first dose of vaccine and the higher (vaccinia-binding) antibody titres achieved after a delayed second dose (and the assumed greater protection this may yield), suggests that the risk threshold favouring a second dose to high-risk subpopulations declines to be only a 3.9-fold higher risk of clinical mpox, or 15-fold higher risk of severe outcomes. These predicted thresholds have wide credibility intervals and rely on a number of assumptions discussed below.

It is important to note that although we discuss in this paper the ratio of cases averted under the different vaccine deployment strategies, this quantity was only possible to calculate here under the assumption of a constant force of infection with time. Given expanding case numbers during an outbreak and the potential impact of public health interventions, risk of infection will likely vary over time. Fortunately, in the supplementary material we show that the same results hold when considering the more complex case of a time varying force of infection (supplementary material). In fact, we show when the force of infection is time variable, a similar quantity to that which we calculate in the main analysis (the ratio of vaccine effectiveness between the two strategies, equation 5) is similarly indicative of the optimal strategy.

Experience with smallpox and Ebola showed that using contact tracing and ring vaccination was a dose-sparing and efficient strategy, albeit with lower vaccine effectiveness due to being administered in a post-exposure prophylaxis setting^22^. Such strategies have the potential to further increase the number of cases averted by controlling the epidemic sooner. However, in our modelling here we have not considered these complementary vaccine deployment strategies. This is largely because there are very limited data available on the risks of infection among contacts, and current estimates for vaccine effectiveness when used as post-exposure prophylaxis have high uncertainty, but may be quite low^5^.

We consider two different methods in our analysis to compare the effectiveness of one- and two-dose regimens. Each of these approaches come with a number of caveats and limitations. In the first approach, we use only the estimates of vaccine effectiveness from meta-analyses of real-world effectiveness studies to compare the ratio of cases averted under the different vaccination strategies (Figs 1E,2C). A strength of this approach is that it does not assume any relationship between antibody titres and protection, and does not consider waning immunity (other than any waning that may have been present in the clinical studies themselves).

However, as a result this approach implicitly assumes vaccine effectiveness does not wane. The second approach involves analysis of the relationship between vaccinia-binding antibody titres and vaccine protection, along with analysis of antibody boosting and waning antibody levels to predict vaccine effectiveness at different times^4^. This is of course significantly less direct than the approach described above and relies on the assumption that vaccinia-binding antibody levels are predictive of vaccine effectiveness against mpox over time, and that antibody titres and immunity will wane in a similar way in the populations at risk of mpox.

The limitations of this modelling approach are outlined in detail in the original study^4^. Further, rather than relying on predictions of the vaccine effectiveness over time from the decay in antibody responses, real world effectiveness data out to 1 to 2 years post MVA-BN vaccination (with either one or two doses) would be very informative. Fortunately, this data is likely available, or soon to be available, in many settings given vaccination during the 2022/2023 clade IIb outbreaks and ongoing clade IIb transmission globally. Further work is clearly needed to validate antibody levels as a predictor of vaccine effectiveness against clinical mpox, against severe mpox, and to investigate antibodies and protection over time in the context of clade I virus.

Both our analysis approaches assume that vaccine supply is low compared to the total susceptible population, such that the deployment strategies do not impact the force of infection. Further, neither approach considers time to epidemic control, where the delay between dose 1 and dose 2 would be influential. Also, both of these methods rely on the results of our meta-analysis of real-world effectiveness studies^4^. There are now three systematic reviews of MVA-BN vaccine effectiveness^4–6^, which give similar estimates of one and two-dose vaccine effectiveness in the context of clade IIb infection. However, the underlying real-world studies included in these meta-analyses each have considerable limitations^4,5^. Further, there is very limited data on vaccine effectiveness against severe mpox infection, and we base our analysis of severe protection on a single meta-analysis of vaccine effectiveness against progression from symptomatic to severe clade IIb infection^5^. Clearly further studies of vaccine effectiveness against clade Ia and Ib and against severe mpox would be valuable to provide data directly relevant to these clades.

Our analysis of vaccine effectiveness against severe mpox relies on the assumption that the vaccine effectiveness estimates we used from real-world effectiveness studies will also apply to the at-risk population. However, vaccine effectiveness may differ for groups at risk of severe mpox such as children, people with HIV, or immunocompromised individuals^23^.

Differences in one- and two-dose effectiveness in such risk groups would change the relative advantage of a one-dose regimen. Recent reports of vaccine effectiveness in people with HIV suggest that this may be similar to the general population^24^. However further studies are clearly needed to understand vaccine immunogenicity and effectiveness across different at-risk populations in order to guide rational vaccine deployment.

Notwithstanding the many limitations of the available data, this analysis suggests that where limited stocks of MVA-BN vaccine are available, deploying them as a single dose regimen to as many people as possible will usually be the most effective strategy to reduce the incidence of mpox. However, providing second doses to high-risk populations may be favoured if the risk-ratio to the general population is sufficiently high. Delaying second doses in the context of limited vaccine supply is not a novel approach. During the COVID-19 pandemic limited supply of the AstraZeneca vaccine in the UK led to increased dosing intervals^25^, and this was accompanied by higher vaccine efficacy than reported with the original (4 week) dosing schedule^26^. These conclusions do not advocate for a change in the recommended vaccine schedule for MVA-BN, or for health authorities to ignore this schedule. Vaccine should be deployed with the intention of giving a second dose at four weeks or as soon as possible. However, limited vaccine availability may at times dictate that allocating limited vaccine supplies as a second dose to those already vaccinated is not an efficient strategy to maximise population protection.

## Methods

### Modelling the averted cases

To compare different vaccine strategies, we calculated the ratio of cases averted between two alternative deployment strategies. Using the estimates for vaccine effectiveness of one- and two-dose regimens reported from the meta-analysis of real-world effectiveness studies, and assuming that effectiveness does not change with time, we previously calculated the ratio of cases averted (RCA) by using the one-dose regimen compared to the two-dose^4^ is given by,

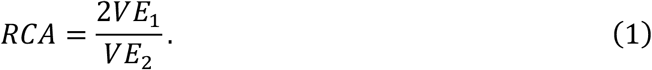

where, *VE*_1_ and *VE*_2_ are the vaccine effectiveness conferred by one and two doses of MVA-BN, respectively.

However, the above approach does not take into account the potential impact of waning immunity over time. We can therefore extend this analysis using our modelled estimates of immune waning and the impact of administering second doses at different times^4^. To model this, we considered a constant force of infection (infection rate), *r*, within the population of concern. We analyse a scenario in which a proportion of individuals has already received the first dose of vaccine, and at some later time more vaccine doses become available that can either be deployed as second doses to those already vaccinated or as first doses to naïve individuals. We assume the number of additional vaccines available (for which a deployment decision is required), *d*, is much less than the vulnerable population, *N*, and that at least *d* people have received a first dose *s* days earlier (see supplementary methods). By assuming the number of vaccines is small, we assume that the vaccine deployment decision does not affect the force of infection.

The cases averted, CA, by a given vaccine strategy compared to a naïve population is then given by,

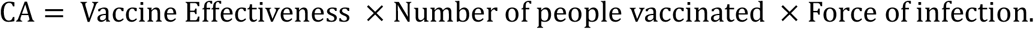

In the case where additional vaccine is deployed in a one-dose regimen to naïve individuals, then the overall protection in the population includes the protection of already vaccinated individuals, plus the protection from the vaccination of naïve subjects. It follows that the expected number of cases averted if the *d* doses are administered to naïve individuals who have not been previously vaccinated is,

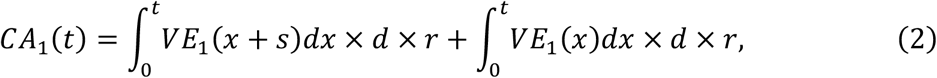

where, *CA*_1_(*t*), is the cumulative number of cases averted by time *t* (in days), and 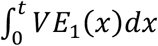 is the cumulative vaccine effectiveness of one dose of MVA-BN over time, *t*, and *r* is the infection rate (i.e. force of infection, see supplement for full derivation). Note that the term *VE*_1_(*x* + *s*) captures that those who were vaccinated with one dose already received this dose *s* days before the current batch of vaccines were deployed, and thus the effectiveness from this dose will have waned for longer than those who have just received their first dose.

Similarly, if these doses are administered as second doses, the number of cases averted by time *t* is given by,

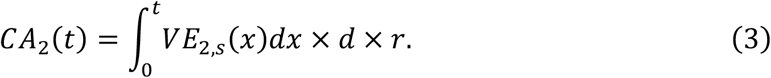

where, *CA*_2_(*t*), is the cumulative number of cases averted by time (*t*) by the two-dose strategy, 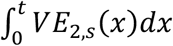 is the cumulative vaccine effectiveness of two doses of MVA-BN over time, *t*, and *s* is the interval between the two doses administered.

For simplicity we assume that the maximum effectiveness is obtained 14 days after vaccination for both one and two doses.

Thus, we can define the ratio of cases averted by time *t* as,

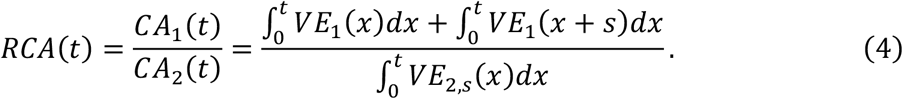

This ratio of cases averted by time *t* is equivalent to the ratio of the average vaccine effectiveness in the population. To calculate the *RCA*, we require estimates of vaccine effectiveness and the waning of immunity. For this we use our estimated vaccine effectiveness, and waning of vaccine effectiveness with time, from our previously published model-based meta-analysis of real world-effectiveness data^4^. In particular, we use the posterior samples of our parameters from our meta-analysis to calculate the ratio in equation 4, and the credible intervals around these ratios. The accuracy for the waning of vaccine effectiveness depends on a number of assumptions from our modelling of the relationship between antibody titres and vaccine effectiveness estimates^4^.

Note that we have employed a model of the ratio of cases averted in this work for ease of interpretation. Importantly, the ratio described in equation 4 only represents the ratio of cases averted under the simplifying assumption of a uniform force of infection over time (supplementary material). However, it should be noted that even in the case where the force of infection is not constant, a very similar ratio, which we call the ratio of vaccine effectiveness in the vaccinatable population (RVE),

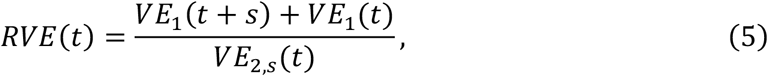

is descriptive of the optimal strategy. As we show in the supplementary material for a time varying force of infection, if this ratio is greater than 1 at all time points, the one dose strategy will be optimal (see derivation in the supplementary methods).

### Incorporating groups with different risk of infection or severe disease

We can further extend this model to consider vaccinating two subpopulations with different risks (different force of infection). In this scenario we consider a high-risk group in which everyone has received a first dose of the vaccine. We then consider a scenario where additional vaccine becomes available and we must decide whether to deploy them as second doses for the already vaccinated high-risk population, or as first doses to a lower-risk population. We can estimate the ‘risk threshold’, which is how much higher the risk of infection must be in the high risk population before targeting the high-risk group with a second dose is favoured. Using only the real-world effectiveness data our meta-analysis^4^, and assuming no waning of immunity, we can estimate that this ratio is,

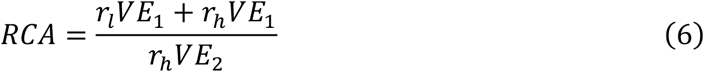

where, *r*_*h*_ and *r*_*l*_ are the rate of infection in the high and low risk groups, respectively. We define the risk threshold, *r*_)_, to be the ratio such that when *r*_*h*_/*r*_*l*_ > *r*_)_, then more cases are averted by administering second doses to the high-risk group. This risk threshold occurs when *RCA* = 1. Thus, more cases will be averted by a one dose strategy, except when

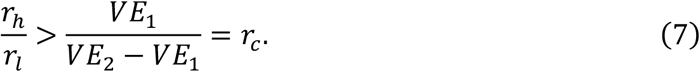

We can also extend this to incorporate the effects of waning immunity and delayed vaccination. In this case, the cases averted by administering a single dose is given by,

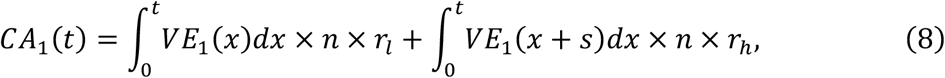

and the cases averted in the two-dose strategy is given by,

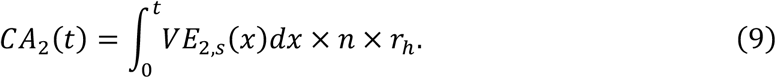

Thus, we can calculate the ratio of cases as,

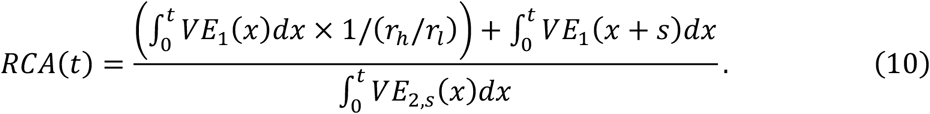

This relationship depends on the infection risk ratio between high risk and lower risk individuals (*r*_*h*_/*r*_*l*_). If the two populations have equal risk (*r*_*h*_/*r*_*l*_ = 1), then we obtain equation 4. Using equation 10, it follows that the risk threshold is given by,

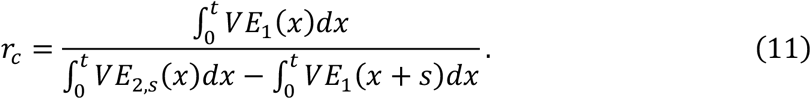

This quantity can be calculated across any time period and in our analysis, and we use a period of two years.

## Data Availability

All data used for analysis and modelling in this present study are available in the original systematic review and meta-analysis reported in Berry et al., Nature Communications 15:3856 (2024) and the accompanying GitHub page

https://github.com/iap-sydney/Mpox_ELISA_Effectiveness_correlates

## Ethical approval

The analysis and modelling of publicly available de-identified clinical trial data was approved under the UNSW Sydney Human Research Ethics Committee (approval HC200242).

