## Supplementary Material for "Optimal deployment of limited vaccine supplies to combat mpox"

### Supplementary Materials

#### The “vaccinatable” population and doses requiring a decision

Assume that we have $D$ vaccine doses for deployment. In order to consider our question of deploying vaccine doses as first or second doses we only wish to model the impact of those fraction of the doses that we actually have a choice to deploy as second doses. I.e. if $k$ individuals have received a first dose, and have not yet been infected, and $k < D$, then only $k$ of our total doses $D$ require a decision of deploying as a first or second dose. On the other hand, if $k \geq D$, then we can choose to deploy all of our doses as second doses. Thus, we only consider the doses of vaccine for which a deployment decision is required, (we call this quantity $d = \min(k, D)$).

Therefore, for the vaccine doses, $d$ we can either deploy them to a group of $d$ people who have already received 1 dose (and who we assume have not been infected, call this population, $N_1 = d$), or a different group of $d$ people who have not received a first dose (call this population, $N_0 = d$). Therefore, we define the “vaccinatable” individuals as these $N_1 + N_0 = 2d$ individuals in our total population to whom we could choose to deploy our $d$ vaccine doses (either as a first dose or a second dose). We assume that the individuals with 1 dose already ($N_1$) received their first dose $s$ weeks before the current vaccine deployment decision of interest and that $s$ is at least 4 weeks (given the MVA-BN vaccine schedule recommends a second dose not earlier than 4 weeks after the first).

#### Generalising the comparison of one and two dose strategies when the force of infection is not constant with time.

Within our analysis we calculate the expected number of cases that occurs within the vaccinatable population ($d$). This analysis used a constant force of infection to model the case numbers and the resulting ratio of cases averted is given by (methods equation 3),

$$RCA(t) = \frac{CA_1(t)}{CA_2(t)} = \frac{\int_0^t VE_1(x)dx + \int_0^t VE_1(x+s)dx}{\int_0^t VE_{2,s}(x)dx}. \quad (1)$$

This quantity translates to the ratio of the average vaccine effectiveness across a group of people (unvaccinated individuals contribute 0 to average effectiveness).

Applying a more sophisticated model, we can model case numbers when the infection rate varies with time. That is, consider an unvaccinated susceptible population, $S_u$, with a varying rate of exposure over time. The number of susceptible unvaccinated individuals over time, $t$, is described by,

$$\frac{dS_u}{dt} = -r(t)S_u, \quad (2)$$

where $r(t)$ is the infection rate at time, $t$. Here we ignore effects of individuals becoming susceptible again (i.e. we assume infected individuals acquire a high level of immunity for the duration of our analysis). Similarly, for a susceptible vaccinated population, $S_v$, we describe this rate of infection being reduced by one minus the vaccine effectiveness (VE), and so the number of vaccinated, susceptible individuals is described by,

$$\frac{dS_v}{dt} = -\big(1 - VE(t)\big)r(t)S_v. \tag{3}$$

Solving, equations (2) and (3) we find that the number of unvaccinated and vaccinated susceptible individuals at a time $t$ are given by,

$$S_u(t) = S_u(0)e^{-\int_0^t r(\lambda)d\lambda} \tag{4}$$

and

$$S_v(t) = S_v(0)e^{-\int_0^t r(\lambda)(1-VE(\lambda))\,d\lambda} = S_v(0)e^{-\int_0^t r(\lambda)d\lambda}e^{\int_0^t r(\lambda)VE(\lambda)\,d\lambda} \tag{5}$$

respectively.

Therefore, the total number of cases that have occurred between time $t = 0$ and time $t$ in the unvaccinated and vaccinated populations are given by,

$$I_u(t) = S_u(0) - S_u(0)e^{-\int_0^t r(\lambda)d\lambda} \tag{6}$$

and

$$I_v(t) = S_v(0) - S_v(0)e^{-\int_0^t r(\lambda)d\lambda}e^{\int_0^t r(\lambda)VE(\lambda)\,d\lambda} \tag{7}$$

respectively.

**The deployment strategies**

We now consider three scenarios

i. Baseline reference scenario

First, as a reference, we consider a scenario of a naïve population. Under this scenario a group of $2d$ naïve individuals are exposed to the force of infection $r(t)$. Thus, from equation (6) above, and noting that in our case we have $S_u(0) = S_v(0) = 2d$, the total number of cases by time $t$ is given by,

$$I_{base}(t) = 2d\left(1 - e^{-\int_0^t r(\lambda)d\lambda}\right) \tag{8}$$

ii. Two dose scenario

Under this scenario, the doses $d$ are deployed to the group $N_1$ as second doses. This provides these individuals with the VE of two doses, but provides individuals in the group $N_0$ with no protection. The second dose is administered at the time of the decision point $t = 0$ and the vaccine effectiveness of this group is given by $VE_{2,s}(t)$, where $s$ is the space between the first and second dose (since VE is influenced by dose spacing). Thus, from equations (6) and (7) the total number of cases that will occur by time $t$ under this scenario is given by,

$$I_{2dose}(t) = d\left(2 - e^{-\int_0^t r(\lambda)d\lambda} - e^{-\int_0^t r(\lambda)(1-VE_{2,s}(\lambda))\,d\lambda}\right) \tag{9}$$

The number of cases averted in the two dose scenario can be calculated by comparing the total number of cases under this scenario with the number of cases under the baseline scenario. Therefore the number of cases averted under the two dose scenario, $CA_2(t)$, is given by:

$$CA_2(t) = I_{base}(t) - I_{2dose}(t) = d\left(e^{-\int_0^t r(\lambda)(1-VE_{2,s}(\lambda))\,d\lambda} - e^{-\int_0^t r(\lambda)d\lambda}\right) \tag{10}$$

iii. One dose scenario

Finally, we consider the scenario where the $d$ doses are deployed as first doses to the $N_0$ group. This leaves the $N_1$ group with the waning protection of their first dose given $s + t$ earlier. From equation (7) it follows that the total number of cases by time $t$ under this scenario is given by,

$$I_{1dose}(t) = d\left(2 - e^{-\int_0^t r(\lambda)(1-VE_1(\lambda))\,d\lambda} - e^{-\int_0^t r(\lambda)(1-VE_1(\lambda+s)\,)d\lambda}\right) \tag{11}$$

The number of cases averted in the one dose scenario can be calculated by comparing the total number of cases under this scenario with the number of cases under the baseline scenario. Therefore the number of cases averted under the one dose scenario, $CA_1(t)$, is given by:

$$\begin{aligned} CA_1(t) &= I_{base}(t) - I_{1dose}(t) \\ &= d\left(e^{-\int_0^t r(\lambda)(1-VE_1(\lambda))\,d\lambda} + e^{-\int_0^t r(\lambda)(1-VE_1(\lambda+s)\,)d\lambda} \right. \\ &\quad \left. - 2e^{-\int_0^t r(\lambda)d\lambda}\right). \end{aligned} \tag{12}$$

Under the above scenarios we have calculated the cases averted using either the one or two dose vaccination strategy. To simplify these relationships further, we now make first order approximation of the exponential terms in equations (10) and (12). This first order approximation assumes that the total force of infection over the relevant time interval is small (in an unvaccinated cohort is small – i.e. $\int_0^t r(\lambda)\,d\lambda \ll 1$, which is akin to an assumption of rare events – of note, since VE is between 0 and 1, it follows that $\int_0^t r(\lambda)VE(\lambda)\,d\lambda < \int_0^t r(\lambda)\,d\lambda \ll 1$). Under this assumption second order and larger terms in the Taylor expansion of an exponential function, given by,

$$e^x = 1 + \mathrm{x} + \frac{\mathrm{x}^2}{2!} + \frac{\mathrm{x}^3}{3!} + \cdots$$

will be very small and thus, we assume,

$$e^{-\int_0^t r(\lambda)(1-VE_{2,s}(\lambda)\,)d\lambda} \approx 1 - \int_0^t r(\lambda)\left(1 - VE_{2,s}(\lambda)\right)\,d\lambda$$

$$e^{-\int_0^t r(\lambda) VE_1(\lambda+s)\, d\lambda} \approx 1 - \int_0^t r(\lambda)(1 - VE_1(\lambda+s))\, d\lambda$$

$$e^{-\int_0^t r(\lambda) VE_1(\lambda)\, d\lambda} \approx 1 - \int_0^t r(\lambda)(1 - VE_1(\lambda))\, d\lambda$$

and

$$e^{-\int_0^t r(\lambda)\, d\lambda} \approx 1 - \int_0^t r(\lambda) d\lambda.$$

Substituting these approximations into equations (10) and (12) gives,

$$CA_2(t) = d\left(\int_0^t r(\lambda)\left(VE_{2,s}(\lambda)\right)\, d\lambda\right) \tag{13}$$

and

$$CA_1(t) = d\left(\int_0^t r(\lambda)\big(VE_1(\lambda+s)\big) d\lambda + \int_0^t r(\lambda)(VE_1(\lambda))\, d\lambda\right). \tag{14}$$

It is clear from the above that the ratio of cases averted (equations 13 and 14) will not be independent of the force of infection, as was the case when the force of infection was constant (main text). However, instead we can look at the difference in cases averted in these two cases,

$$\begin{aligned} CA_1(t) - CA_2(t) &= d\left(\int_0^t r(\lambda)\big(VE_1(\lambda+s)\big) d\lambda + \int_0^t r(\lambda)(VE_1(\lambda))\, d\lambda \right. \\ &\left. - \int_0^t r(\lambda)\left(VE_{2,s}(\lambda)\right)\, d\lambda\right). \end{aligned} \tag{15}$$

Therefore, it follows that for the cases averted by the one dose strategy to be greater than the two dose strategy we require that,

$$\begin{aligned} CA_1(t) - CA_2(t) &= d\left(\int_0^t r(\lambda) VE_1(\lambda)\, d\lambda + \int_0^t r(\lambda) VE_1(\lambda+s)\, d\lambda \right. \\ &\left. - \int_0^t r(\lambda) VE_{2,s}(\lambda)\, d\lambda\right) > 0 \end{aligned} \tag{16}$$

and this can only be true when

$$\left(\int_0^t r(\lambda) VE_1(\lambda)\, d\lambda + \int_0^t r(\lambda) VE_1(\lambda+s)\, d\lambda - \int_0^t r(\lambda) VE_{2,s}(\lambda)\, d\lambda\right) > 0, \tag{17}$$

since $d > 0$. Combining these integrals, we see that we require that

$$\left(\int_0^t r(\lambda)\left(VE_1(\lambda)\ + VE_1(\lambda+s) - VE_{2,s}(\lambda)\right)d\lambda\right) > 0. \tag{18}$$

We notice that under the strict condition that $VE_1(\lambda)\ + VE_1(\lambda+s) - VE_{2,s}(\lambda) > 0$ for all $0 < \lambda < t$, equation (18) will hold true. Thus, rearranging this expression, we see that the one dose strategy will avert more cases than the 2 dose strategy if the ratio (which we call the vaccine effectiveness of the vaccinatable population, RVE),

$$RVE(t) = \frac{VE_1(t) + VE_1(t+s)}{VE_{2,s}(t)} > 1, \tag{19}$$

at all time-points in our time interval of interest (2 years in the main text).

In the same way the cases averted by the two dose strategy will be greater than the one dose when the ratio $RVE(t) < 1$ at all time-points.

Importantly, this ratio is greater than 1 at all time points for the MVA-BN vaccine regimens we are considering (figure S1). Therefore, our result that more cases will be averted by administering limited vaccine doses as first doses to as many individuals as possible rather than as second doses to those already vaccinated in the main text holds independently of assumptions of a constant force of infection.

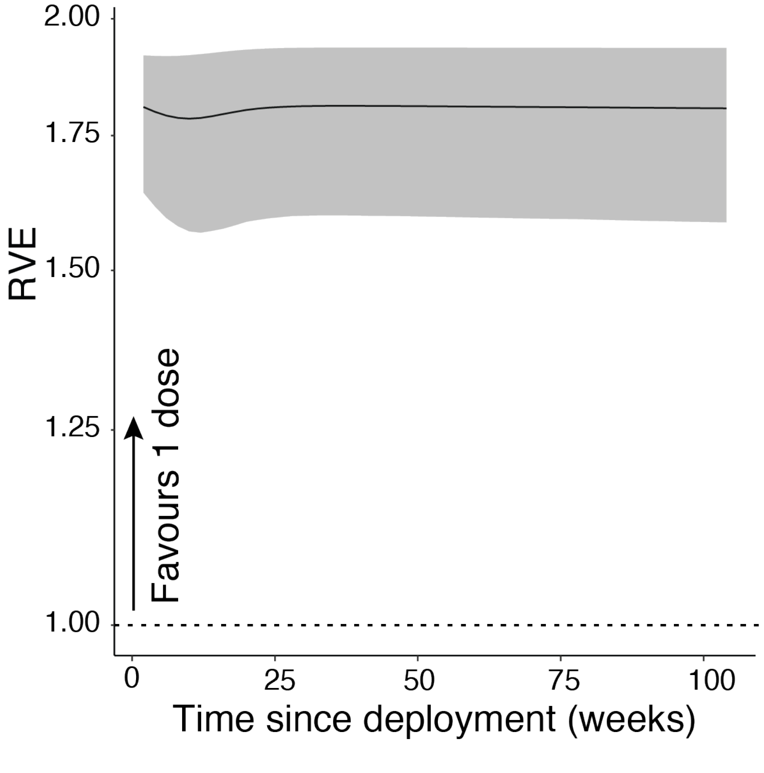


**Figure S1:** The ratio of vaccine effectiveness at each time point, $RVE(t)$, from equation (19) when the space between doses is, $s$, 4 weeks. Since this ratio is always greater than 1 over the 2 year period of interest, it follows (as described above) that a one-dose vaccination strategy will avert more cases than a two dose strategy, given limited vaccine supply. Note that the same result holds whether the spacing of doses is 6 months or 1 year (not shown).

### Considering a time varying force of infection along with different risks of infection or severe outcomes

Finally, we consider the case of a time varying force of infection and two groups with different force of infection. We assume the force of infection in the high risk group is given by $r_h(t) = r_h h(t)$ and the risk in the low risk group is given by $r_l(t) = r_l h(t)$, where $h(t)$ is the time varying component of the force of infection. Under this assumption the relative risk between the two groups is constant over time, but the absolute force of infection for each group is time varying. Under this assumption we see that,

$$I_{2dose}(t) = d\left(2 - e^{-r_l \int_0^t h(\lambda)d\lambda} - e^{-r_h \int_0^t h(\lambda)d\lambda} e^{r_h \int_0^t h(\lambda)VE_{2,s}(\lambda)\, d\lambda}\right) \tag{20}$$

and

$$I_{1dose}(t) = d\left(2 - e^{-r_l \int_0^t h(\lambda)d\lambda} e^{r_l \int_0^t h(\lambda)VE_1(\lambda)\, d\lambda} - e^{-r_h \int_0^t h(\lambda)d\lambda} e^{r_h \int_0^t h(\lambda)VE_1(\lambda+s)\, d\lambda}\right) \tag{21}$$

The difference in cases averted between these two strategies is given by,

$$\begin{aligned} CA_1(t) - CA_2(t) &= d\left(e^{-r_l \int_0^t h(\lambda)d\lambda} + e^{-r_h \int_0^t h(\lambda)d\lambda} e^{r_h \int_0^t h(\lambda)VE_{2,s}(\lambda)\, d\lambda}\right. \\ &- e^{-r_l \int_0^t h(\lambda)d\lambda} e^{r_l \int_0^t h(\lambda)VE_1(\lambda)\, d\lambda} \\ &\left.- e^{-r_h \int_0^t h(\lambda)d\lambda} e^{r_h \int_0^t h(\lambda)VE_1(\lambda+s)\, d\lambda}\right) \end{aligned} \tag{22}$$

Taking a first order approximation, as above, and expanding and simplifying we find that

$$\begin{aligned} I_{1dose}(t) - I_{2dose}(t) &\approx d\left(r_l \int_0^t h(\lambda)VE_1(\lambda)\, d\lambda + r_h \int_0^t h(\lambda)VE_1(\lambda+s)\, d\lambda\right. \\ &\left.- r_h \int_0^t h(\lambda)VE_{2,s}(\lambda)\, d\lambda\right). \end{aligned} \tag{23}$$

Thus, as above $I_{1dose}(t) - I_{2dose}(t) > 0$ only when,

$$\int_0^t h(\lambda)\left(r_l VE_1(\lambda) + r_h VE_1(\lambda+s) - r_h VE_{2,s}(\lambda)\right)\, d\lambda > 0. \tag{24}$$

Equation (24) will be satisfied whenever

$$r_l VE_1(\lambda) + r_h VE_1(\lambda+s) - r_h VE_{2,s}(\lambda) > 0 \tag{24}$$

for all $0 < \lambda < t$.

Thus, the one dose strategy will avert more cases than the 2 dose strategy whenever,

$$\frac{r_h}{r_l} > \frac{VE_1(t)}{VE_{2,s}(t) - VE_1(t+s)} \tag{25}$$

for time points in our analysis. From this we see that in the case of a time varying force of infection, we can define a critical relative risk threshold between high risk and low risk individuals, such that when the ratio in equation 25 is satisfied, we can be confident the one dose-strategy is superior. This proof leaves open the possibility that even in some cases where equation (25) is not satisfied, the one-dose strategy may be optimal.
